# A novel taxonomy to assess dressing activity in chronic stroke

**DOI:** 10.1101/2023.10.04.23295488

**Authors:** Emily E. Fokas, Zuha Ahmed, Avinash R. Parnandi, Anita Venkatesan, Natasha G. Pandit, Dawn M. Nilsen, Heidi M. Schambra

**Affiliations:** Department of Neurology, New York University Grossman School of Medicine; Department of Medicine, New York University Grossman School of Medicine; Department of Rehabilitation and Regenerative Medicine, Columbia University; Programs in Occupational Therapy, Columbia University; Department of Rehabilitation Medicine, New York University Grossman School of Medicine

## Abstract

Upper-body dressing (UBD) is a key aspect of motor rehabilitation after stroke, but most individuals with stroke require long-term dressing assistance. Having a measurement approach that captures the quantity and quality of dressing movements during training could support more targeted strategies. As the basis of an approach, we modified our previously developed motion taxonomy, which categorizes elemental motions into classes of functional primitives (e.g. *reaches, transports, stabilizations*). Three expert coders examined videos of two healthy subjects performing dressing tasks, and expanded the taxonomy to account for the unique arm and trunk motions of UBD. An expert and a trained coder then applied the expanded taxonomy to dressing videos of five chronic stroke subjects. We examined the interrater reliability (IRR) for classifying primitives. Using the expanded taxonomy, IRR for identifying primitives in UBD was overall low (k = 0.52) but varied by primitive class: IRR was moderate for *reach* (k = 0.75), *transport* (k = 0.63), and *idle* (k = 0.68), lower for *reposition* (k = 0.58), and negligible for *stabilization* (k = -0.02). IRR increased with increasing UE-FMA score (ρ=1, p<0.0001), indicating that the reliability of primitive classification improved with less impaired movement. With additional modification, the expanded taxonomy could support the measurement of training doses and impaired motion during dressing activities.

## Introduction

Upper-body dressing (UBD) is a particularly intimate activity of daily living (ADL) and is a key aspect of motor rehabilitation after stroke, but 36% of patients continue to require dressing assistance two years after their stroke (Edmans et al., 1991). Measurement approaches that capture the quantity and quality of dressing movements are currently lacking, and may lead to suboptimal training that contributes to chronic dependency. Previous UBD assessments, such as the Nottingham Stroke Dressing Assessment (Walker & Lincoln, 1990) and the UBD-Scale (Suzuki et al., 2008), score patients by the level of assistance needed. Although useful predictors of UBD recovery, these scales do not measure motion quantity or abnormality in dressing activities. Taxonomies that decompose dressing into component actions such as ‘getting necessary clothes from closets and drawers’ and ‘dressing upper trunk’ (Tornquist & Sonn, 1994) do not support measurement of training quantity in UBD because their constituent motions can vary greatly in type and number.

We previously created a functional motion taxonomy that breaks down upper extremity (UE) activities into five classes of motion elements, called functional primitives: *reach, reposition, transport, stabilization*, and *idle* (Schambra et al., 2019). Functional primitives are strung together to complete basic and instrumental ADLs (IADLs) and their identification serves as the basis for measuring movement repetitions (Parnandi et al., 2022) and movement quality (Parnandi et al., 2023). However, the functional motion taxonomy has not been applied to UE motions used in UBD. ADL/IADLs commonly require the UEs to engage a rigid object in peri-personal space in front of the individual. Some components of UBD, like removing a buttoned shirt from a hanger and unfolding a t-shirt, similarly operate in the peri-personal space. However, dressing is unique in that it also requires the UE to engage a fluid object—clothing—in the personal space that envelops the UE, and this object must be managed above and behind the individual (Ramisa et al., 2014). Thus, we examined whether an expanded version of our functional motion taxonomy could be used to reliably classify UBD primitives. We also examined whether the degree of UE motor impairment affected the reliability of classification. If this expanded taxonomy can successfully decompose UBD into recognizable units of functional motion, it could serve the quantitation of UBD training and the assessment of UBD motion quality.

## Methods

### Subjects

Human experimentation was approved by the local institutional review board, and informed consent from subjects was obtained. We studied the video recordings of two healthy and five stroke subjects that were previously obtained in another study (Parnandi et al., 2022). All subjects were right-handed and stroke subjects had left hemisphere stroke at least 6 months before with right UE weakness (Medical Research Council score <5/5 in at least one muscle group).

### Study Design

Subjects performed one trial each of donning and doffing a t-shirt, a buttoned shirt, and a zippered jacket. Articles of clothing were placed at their right (in stroke subjects, paretic) side. For each trial, we instructed subjects to unfold the clothing, put it on, take it off, and return it to the investigator; no other instructions were given. Upper body motion was recorded with two Ninox 125 video cameras (60 frames/second, 1088 × 740 resolution) positioned approximately two meters orthogonally to the subject. UE motor impairment was also measured with the UE Fugl-Meyer Assessment (UE-FMA) (Fugl-Meyer et al., 1975).

To define the characteristics of UE primitives used in UBD, three experts in the standard functional motion taxonomy (EF, AP, HS) examined the recordings from the two healthy subjects. Through consensus process, descriptions of standard primitives were modified to account for motion variants unique to UBD (Table 1). Unlike standard primitives in the peri-personal space, dressing-specific primitives generally used more sinuous UE motion and truncal motion, had changing degrees of contact with the target object (clothing), and object contact was typically made with the entire UE (Figure 1).

**Table 1.**
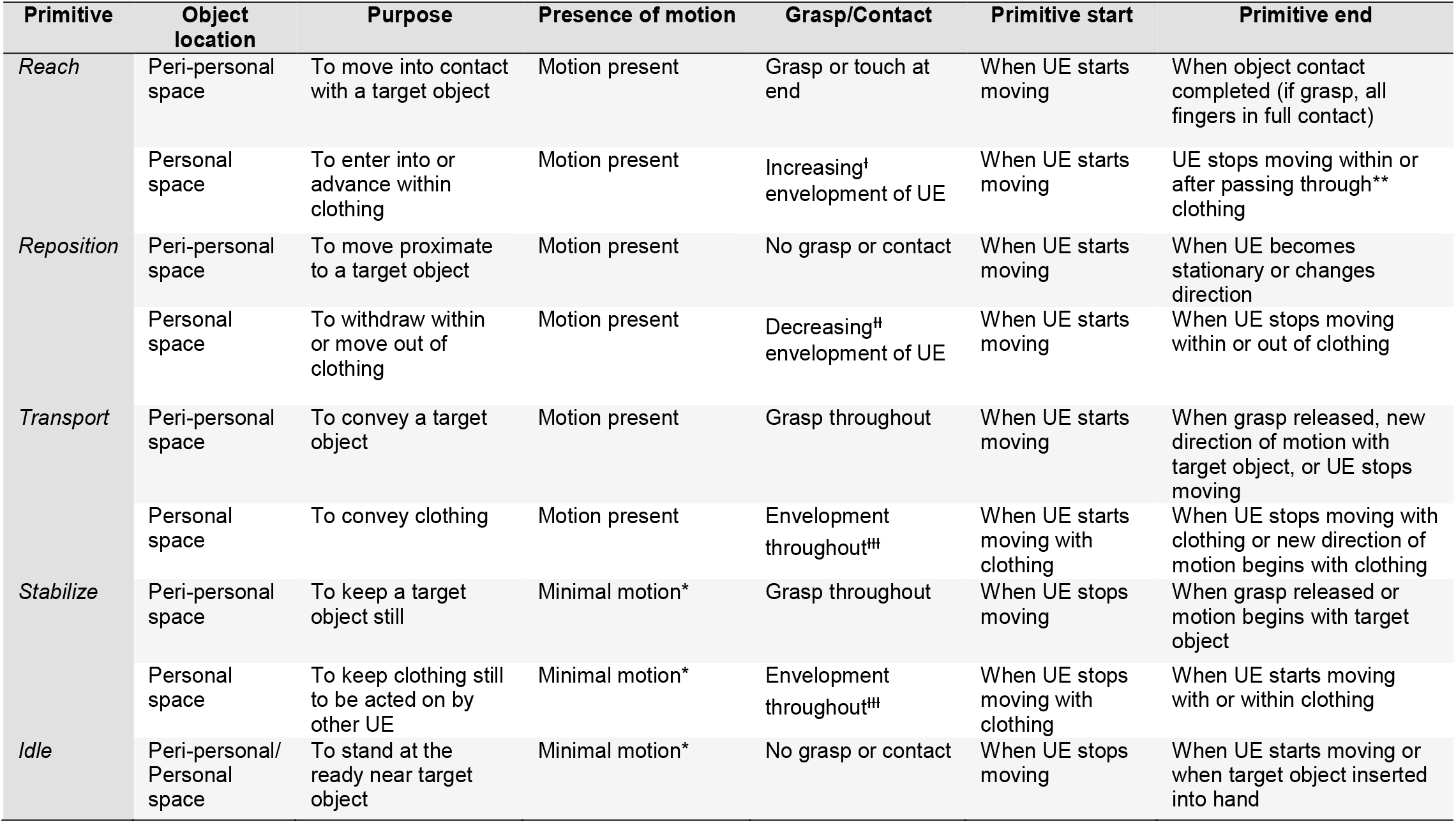
Expanded functional motion taxonomy with upper-body dressing (UBD) primitives. Shown are comparisons between our existing functional motion taxonomy ^5^, which characterizes primitives in feeding, grooming, and IADLs, versus the expanded primitive types seen in UBD. In IADLs, the UE typically engages objects in the peri-personal space in front of the individual. UBD, conversely, requires the UE to engage the object—clothing—in both the peri-personal and personal space that envelops the UE. The purpose of the primitive is analogous between the existing and proposed UBD primitives. The presence of motion is the translation of the UE in space with respect to the pelvis. Unlike IADLs, in which object contact is typically hand-centric, object contact in UBD primitives typically involves the envelopment of the entire UE by clothing. The primitive start and primitive end were operationalized for data segmentation purposes, and are expanded for UBD to account for enveloping contact by clothing. *UE location remains largely unchanged for ≥ 50 ms; subtle drift in the UE is disregarded. **UE may have exited portion of clothing (e.g. sleeve) but motion ends after exiting. ^ᶧ^Clothing may be fully on UE early in motion. ^ᶧᶧ^Clothing may be fully off UE early in motion. ^ᶧᶧᶧ^Clothing may be shifting location on the UE.

| Primitive | Object location | Purpose | Presence of motion | Grasp/Contact | Primitive start | Primitive end |
| --- | --- | --- | --- | --- | --- | --- |
| *Reach* | Peri-personal space | To move into contact with a target object | Motion present | Grasp or touch at end | When UE starts moving | When object contact completed (if grasp, all fingers in full contact) |
|  | Personal space | To enter into or advance within clothing | Motion present | Increasing<sup>†</sup> envelopment of UE | When UE starts moving | UE stops moving within or after passing through<sup>**</sup> clothing |
| *Reposition* | Peri-personal space | To move proximate to a target object | Motion present | No grasp or contact | When UE starts moving | When UE becomes stationary or changes direction |
|  | Personal space | To withdraw within or move out of clothing | Motion present | Decreasing<sup>#</sup> envelopment of UE | When UE starts moving | When UE stops moving within or out of clothing |
| *Transport* | Peri-personal space | To convey a target object | Motion present | Grasp throughout | When UE starts moving | When grasp released, new direction of motion with target object, or UE stops moving |
|  | Personal space | To convey clothing | Motion present | Envelopment throughout<sup>‡‡</sup> | When UE starts moving with clothing | When UE stops moving with clothing or new direction of motion begins with clothing |
| *Stabilize* | Peri-personal space | To keep a target object still | Minimal motion* | Grasp throughout | When UE stops moving | When grasp released or motion begins with target object |
|  | Personal space | To keep clothing still to be acted on by other UE | Minimal motion* | Envelopment throughout<sup>‡‡</sup> | When UE stops moving with clothing | When UE starts moving with or within clothing |
| *Idle* | Peri-personal/ Personal space | To stand at the ready near target object | Minimal motion* | No grasp or contact | When UE stops moving | When UE starts moving or when target object inserted into hand |

**Figure 1.**
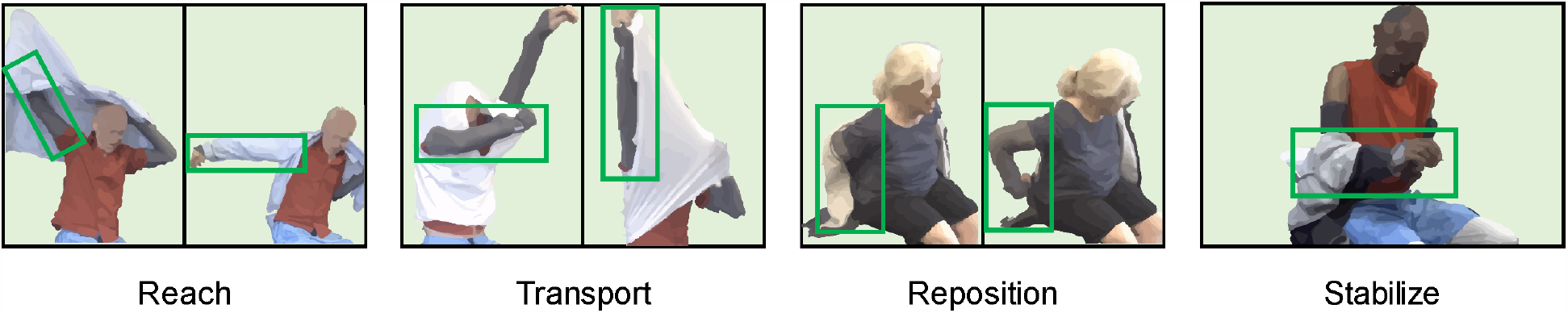
Identification of motion primitives in dressing tasks. Examples of functional motion primitives identified during t-shirt, buttoned shirt, and zippered jacket task performed by the right UE (highlighted in green).

Once UBD primitives were operationalized, a trained coder (ZA) and an expert (EF) reviewed the recordings of the five stroke subjects. Each coder labeled the onset and offset of primitives performed by the paretic UE.

### Analyses

We compared labeled primitives between trained coder and expert. We examined interrater reliability (IRR) with Cohen’s Kappa (McHugh, 2012) for the dressing tasks overall, for each primitive class, and for each subject. We also examined the reliability of primitive classification across the range of UE impairment. We used Spearman’s correlation to examine whether subject-level IRRs related to their UE-FMA scores. We performed all analyses in JMP Pro 16 (SAS), with the significance of uncorrected p-values set at α=0.05. Data are reported as mean ± standard deviation.

## Results

Our study sample included two healthy subjects (1 male, age 63.4 ± 8.8 years, UE-FMA 65.5 ± 0.7 points) and five stroke subjects (2 male, age 56.4 ± 9.1 years, UE-FMA 47.8 ± 10.5 points, time post-stroke 5.4 ± 6.2 years). In the stroke subjects, the three dressing tasks generated 125.4 ± 18.8 primitives, of which 23.8 ± 7.2 were UBD-specific.

The overall IRR for classifying primitives in the dressing activities was low on average, with k = 0.52. Examined by primitive class, IRR was moderate for *reach* (k = 0.75), *transport* (k = 0.63), and *idle* (k = 0.68), lower for *reposition* (k = 0.58), and negligible for *stabilization* (k = -0.02). Compared to the expert, the trained coder misclassified 50% of *stabilizations* as *transports* and 25% as *idles*. Examined by impairment level, IRR increased with increasing UE-FMA score (ρ=1, p<0.0001; Fig. 2), indicating that the reliability of primitive classification improved with less impaired movement.

**Figure 2.**
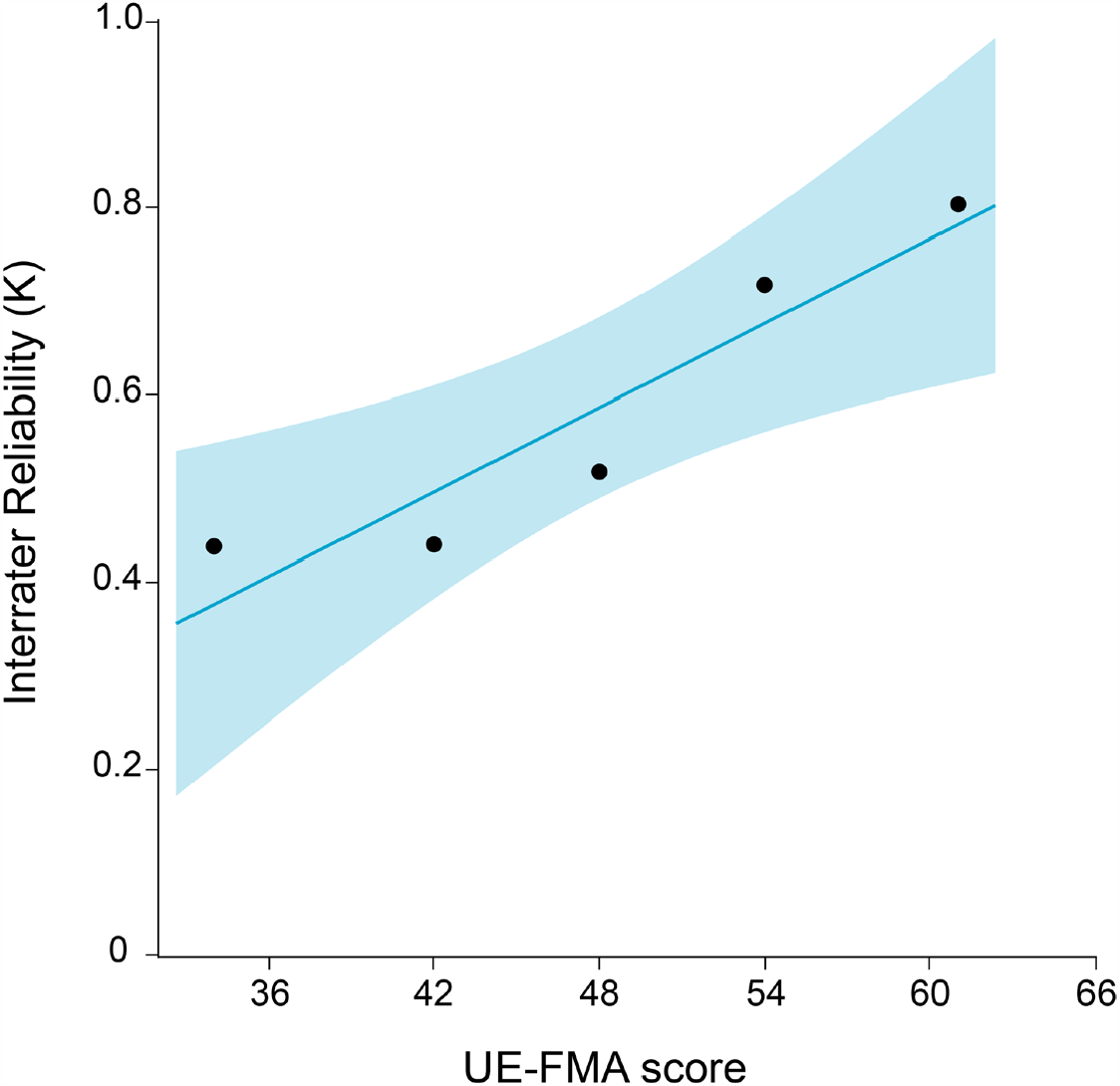
Relationship between motion primitive identification and motor impairment in stroke. Data shown are the correlation between Cohen’s Kappa and UE-FMA scores. Higher Cohen’s Kappa values were significantly related to higher UE-FMA scores (less motor impairment; p<0.0001). Note that data are shown on a continuous scale for visualization, but ranked correlation analysis was performed.

## Discussion

The proposed taxonomy, expanded to classify primitives in UBD, was effective at breaking down dressing into component motions. This taxonomy is unique from existing dressing taxonomies (Tornquist & Sonn, 1994) and clinical scales (Suzuki et al., 2008; Walker & Lincoln, 1990). The three dressing tasks generated a high number of primitives compared to other ADLs, such as drinking a bottle of water and applying deodorant, where counts generally range from 74-80 primitives (Schambra et al., 2019). The high number of primitives generated highlights the value of dressing ADLs in dose-based training, and underscores the need for a measurement taxonomy of this kind.

The defined phenotypes of UBD primitives supported moderate interrater reliability for most primitives except *stabilization*. The general UE stillness associated with this primitive likely contributed to its confusion for an *idle*, whereas a subtle UE configuration drift permitted in this primitive may have contributed to its confusion for a clothing *transport*. Further refinement of the taxonomy is required to help distinguish between these primitive types.

The expanded taxonomy was reliable in mildly impaired subjects, but was less reliable in moderately impaired subjects. This poorer reliability may be due in part to the diminished UE movement extents and the increased use of truncal motions, especially for reaches and transports, in the more impaired subjects. These compensatory motions rendered primitives less distinct and more challenging to classify. The consideration of truncal motion in the definition of UBD primitives may be useful for their classification in moderately impaired subjects, an area for continued work.

### Study Limitations

Our study was limited to a small sample of moderately and mildly impaired stroke subjects, and thus it is unclear how the taxonomy would perform in severely impaired subjects (UE-FMA <25). Future work will explore the application of the taxonomy at this impairment level.

## Conclusions

In summary, the identification of motion-based UBD primitives could be used to measure training dose and assess motion abnormality in dressing activities performed by mildly impaired stroke subjects.

## Data Availability

All data produced in the present study are available upon reasonable request to the authors.

